# Risk and severity of SARS-CoV-2 reinfections during 2020-2022 in Vojvodina, Serbia: a population-level study

**DOI:** 10.1101/2022.04.08.22273571

**Authors:** Snežana Medić, Cleo Anastassopoulou, Zagorka Lozanov-Crvenković, Vladimir Vuković, Nataša Dragnić, Vladimir Petrović, Mioljub Ristić, Tatjana Pustahija, Zoran Gojković, Athanasios Tsakris, John P. A. Ioannidis

## Abstract

**Background:** Data on the rate and severity of SARS-CoV-2 reinfections in real-world settings are scarce and the effects of vaccine boosters on reinfection risk are unknown.

**Methods:** In a retrospective cohort study, registered SARS-CoV-2 laboratory-confirmed Vojvodina residents, between March 6, 2020 and October 31, 2021, were followed for reinfection ≥90 days after primary infection. Data were censored at the end of follow-up (January 31, 2022) or death. The reinfection risk was visualized with Kaplan-Meier plots. To examine the protective effect of vaccination, the subset of individuals with primary infection in 2020 (March 6-December 31) were matched (1:2) with controls without reinfection.

**Findings:** Until January 31, 2022, 13,792 reinfections were recorded among 251,104 COVID-19 primary infections (5.49%). Most reinfections (86.8%) were recorded in January 2022. Reinfections were mostly mild (99.2%). Hospitalizations were uncommon (1.8% *vs*. 3.70% in primary infection) and COVID-19 deaths were very rare (n=20, case fatality rate 0.15%). The overall incidence rate of reinfections was 5.99 (95% CI 5.89-6.09) per 1,000 person-months. The reinfection risk was estimated as 0.76% at six months, 1.36% at nine months, 4.96% at 12 months, 16.7% at 15 months, and 18.9% at 18 months. Unvaccinated (OR=1.23; 95%CI=1.14-1.33), incompletely (OR=1.33; 95%CI=1.08-1.64) or completely vaccinated (OR=1.50; 95%CI=1.37-1.63), were modestly more likely to be reinfected compared with recipients of a third (booster) vaccine dose.

**Interpretation:** SARS-CoV-2 reinfections were uncommon until the end of 2021 but became common with the advent of Omicron. Very few reinfections were severe. Boosters may modestly reduce reinfection risk.

## Introduction

Reinfections with SARS-CoV-2 are an important aspect of COVID-19 and its potential transition to endemicity.^1-3^ Current challenges include the absence of widespread genomic surveillance, the durability of immunity after primary infection, and the need, timing and variant target of booster doses.^4^

Reinfections, commonly defined by a positive RT-PCR test ≥90 days from the first episode, were rare (absolute rate 0-1·1%) before COVID-19 vaccines were available.^5-7^ As mass vaccination proceeded and longer follow-up accrued, reinfection parameters, particularly pertaining to severity, are important to evaluate. Recent studies showed that reinfection is less likely in vaccinated individuals who have recovered from a previous infection.^8,9^

This study aimed to assess the SARS-CoV-2 reinfection rate and associated severity in the population of the Autonomous Province of Vojvodina, which comprises almost a third of Serbia’s population. A secondary aim was to determine the potential reinfection prevention benefit of vaccination in persons who have recovered from an initial episode.

## Materials and methods

### Study setting, objectives, and data collection

The study was conducted at the Institute of Public Health of Vojvodina (IPHV), an integrated health institution responsible for the oversight of disease control and prevention for ≈1·9 million Vojvodina inhabitants. Reinfection was defined as the detection of SARS-CoV-2 RNA or antigen in nasopharyngeal swab specimens after ≥90 days from the first episode (primary infection), regardless of the presence of symptoms.

Our primary objective was to determine the risk of reinfection and associated severity in Vojvodina residents with primary infection (first positive SARS-CoV-2 test) between March 6, 2020 and October 31, 2021. Data were censored at the end of follow-up (January 31, 2022) or death. A secondary objective was to examine whether vaccination protected from reinfection. For this purpose, the risk of reinfection was estimated for the subset of participants with primary infection in 2020 (March 6– December 31), before vaccines were available.

Socio-demographic and epidemiological data were retrieved from the IPHV surveillance database which contains data for all registered COVID-19 cases in Vojvodina from the beginning of the pandemic (March 6, 2020). All cases were residents of Vojvodina. Data originated from epidemiological questionnaires and mandatory notification forms. The minimum set of data for each laboratory-confirmed case included demographics, occupation, date of symptom(s) onset and list of symptoms, severity of COVID-19 disease, date and type of positive laboratory diagnostic test, date of case registration, number of comorbidities, hospitalization status (hospitalized *vs*. non-hospitalized), disease outcome with a specified date (active/recovered/death outcome) and COVID-19 vaccination status (type of vaccine, number of doses and date(s) of vaccination(s)). Regarding disease severity, for surveillance purposes, COVID-19 cases were classified into three groups: 1) mild, if there were no symptoms or COVID-19-related symptoms were present but without confirmed pneumonia by chest imaging; 2) severe, if COVID-19 pneumonia was confirmed by chest imaging; and 3) critical, if COVID-19 pneumonia required mechanical ventilation and/or admission to the intensive care unit.

### Laboratory testing

For the purpose of SARS-CoV-2 detection, posterior nasopharingeal swab samples were collected as part of case-based surveillance protocol for COVID-19 or due to other indications, regardless of reason for testing. Different semi-quantitative RT-PCR tests were used, depending on availability; results were interpretated in accordance with the manufacturer’s instructions.^10-13^ Antigen rapid diagnostic tests (Ag-RDT), for the qualitative detection of SARS-CoV-2 antigens in nasopharyngeal swabs, were introduced in November 2020. Patients with COVID-19-related symptoms within five days of symptom(s) onset were tested using the STANDARD Q COVID-19 Ag Test (SD Biosensor, Gyeonggi-do, South Korea).^14,15^ Patients with COVID-19-related symptoms and negative Ag-RDT results were subsequently tested by RT-PCR in a repeated nasopharyngeal swab for a definitive diagnosis.^16^ A COVID-19 laboratory-confirmed case was defined as a subject with detected SARS-CoV-2 RNA or antigen in a nasopharyngeal swab by a positive RT-PCR test or Ag-RDT, respectively.

### Assessment of the protective effect of vaccination towards reinfection Eligibility criteria and case/control definitions

*The study case* was a resident of Vojvodina, with laboratory-confirmed SARS-CoV-2 infection reported in the period March 6, 2020 to December 31, 2020, with confirmed reinfection from January 1, 2021 to January 31, 2022. *The study control* was a resident of Vojvodina with a laboratory-confirmed SARS-CoV-2 infection reported in the period March 6 to December 31, 2020 and without laboratory confirmation of reinfection from January 1, 2021 to January 31, 2022.

#### Exclusion criteria

To examine the relationship between vaccination and SARS-CoV-2 reinfections, 25 detected reinfection cases in 2020, when vaccination was still unavailable, were excluded. For the same reason, the status of all study cases and controls was checked in the provincial mortality database and in case of death outcome (regardless of cause) before January 31, 2022, were excluded from analysis (12 study cases and 2,784 controls). Since children did not have the same chances for vaccination as adults (recommendations for vaccination of children aged 12-17 years were issued in June 2021), 2051 children and adolescents aged <18 years were excluded from analysis.

### Matching of study cases and controls

The IPHV electronic database was searched to assess the SARS-CoV-2 reinfection rate in the study period and to identify study cases and controls, according to eligibility criteria and case definitions. To document reinfection, we used the Unique Master Citizen Number (ID number) that accompanies all reported cases. For 5·6% of persons for whom the ID number was missing, the search was performed by patient name, surname, date of birth and residence. Matching of study cases and controls was done in a ratio of 1:2. Each study case was paired individually with a control in relation to gender, date of initial SARS-CoV-2 positive test (±10 days) and corresponding age (±3 years). Random selection was applied if several controls corresponded to a study case by using a random number generator. Of 7,083 study-cases ≥18 years, 12 (0·2%) were excluded due to unsuccessful matching in relation to applied criteria.

### Classification of study cases and controls by vaccination status

The following definitions were used: Unvaccinated status was considered if no vaccine dose was received or if a single (or the first of a two-dose scheme) dose was received ≤14 days before the reinfection date, which was the date of reinfection symptoms onset (or positive test, in the absence of symptoms). Incomplete (partial) vaccination was considered if a single vaccine dose was received and >14 days had passed since the vaccination day; if the vaccination schedule had not been completed; or if the final dose was given ≤14 days or >6 months before the reinfection date. Fully vaccinated status was considered if two vaccine doses were administered and the final dose was received >14 days and ≤6 months before the reinfection date. Boosted were considered patients who received the third (booster) dose and >7 days had passed since receiving the vaccine before the reinfection date. The same definitions were applied to controls.

### Statistical analysis

Descriptive analysis of socio-demographic, epidemiological and clinical features, including reinfections severity, was conducted. Cochran–Armitage test was used to assess trend of proportions of primary infection that were reinfected in the observed period. We assessed the risk of suspected reinfection, using time-to-event Kaplan-Meier analysis to estimate the reinfection risk during follow-up. Only patients who survived three months after primary infection were considered for analysis (since, by definition, there can be no reinfection risk in the first three months). We also show Kaplan-Meir plots separately for primary infections that occurred in time periods defining each pandemic wave. Each of these curves were plotted for those who were first infected in that time interval. Using the same analysis, we additionally estimated the risk of severe reinfection or worse outcome (severe/critical/death) over time, again starting at three months after primary infection. Pearson’s chi-squared test or Fisher’s exact test, as appropriate, was used to compare differences between groups of categorical set of data. For variables with multiple ordered categories, we used analyses adjusted for trend. Binary logistic regression was applied to assess the association between different factors and reinfection severity. Conditional logistic regression was applied to compare unvaccinated, incomplete and full vaccination with boosted vaccination status as a reference variable among study-cases and controls. McNemar test was used to test difference in proportions between paired data. A p-value <0·005 was considered statistically significant and p-values <0·05 were considered suggestive.^17^ Stata v.16 (College Station, TX: StataCorp LLC. 2019) and TIBCO Statistica™ 14.0.0 (license for University of Novi Sad) were used for matching and statistical analyses.

### Ethics Statement

This case-control study was performed in the frame of national public health surveillance. Sample collection for laboratory diagnosis of COVID-19 formed part of the standard patient management, thus, only oral informed patient consent was required. Access to patients’ data was restricted for employees directly involved in COVID-19 diagnosis, treatment, and reporting. Patients’ data were anonymized before any analysis was conducted. According to the law, no approval by the Ethics Committee for the retrospective analysis of anonymized data is required in Serbia.

## Results

### SARS-CoV-2 reinfections over time

A total of 362,650 COVID-19 cases were registered among the population of Vojvodina from the beginning of the pandemic (March 6, 2020) until January 31, 2022 (Table 1). The cohort included a subset of 251,104 patients with primary infection in the period March 6, 2020-October 31, 2021, of which 13,792 (5·49%) experienced reinfection (Table 2). Before August 2021, reinfections were sporadically registered (rate <1%). Most reinfections (86·77%) were recorded in January 2022, when the reinfection rate abruptly increased up to 15·32%. The highest proportion of primary infections that were reinfected was in October and November 2020 (11·05% and 10·67%, respectively), with decreasing linear trend after October 2020 (SI Fig.1). The average time duration from primary infection to reinfection was 340±101 days (min 90 days–max 662 days).

**Table 1.** **Proportion of patients with SARS-CoV-2 reinfection in relation to overall registered COVID-19 cases in Vojvodina, Serbia, March 6, 2020-January 31, 2022**.

|  | Months | Overall registered COVID-19 cases<sup>†</sup> |  | Proportion of reinfections<sup>§</sup> |  |
| --- | --- | --- | --- | --- | --- |
|  |  | n | % | n | % |
| 2020 | March-June | 1,570 | 0.43 | - | - |
|  | July | 4,597 | 1.27 | 0 | 0 |
|  | August | 1,750 | 0.48 | 0 | 0 |
|  | September | 330 | 0.09 | 0 | 0 |
|  | October | 1,547 | 0.43 | 1 | 0.06 |
|  | November | 30,659 | 8.45 | 9 | 0.03 |
|  | December | 38,392 | 10.59 | 15 | 0.04 |
|  | **Total** | **78,845** | **-** | **25** | **0.03** |
| 2021 | January | 11,441 | 3.15 | 7 | 0.06 |
|  | February | 11,868 | 3.27 | 11 | 0.09 |
|  | March | 30,012 | 8.27 | 47 | 0.16 |
|  | April | 21,929 | 6.05 | 47 | 0.21 |
|  | May | 5,084 | 1.40 | 21 | 0.41 |
|  | June | 586 | 0.16 | 5 | 0.85 |
|  | July | 860 | 0.24 | 8 | 0.93 |
|  | August | 7,029 | 1.94 | 80 | 1.14 |
|  | September | 35,515 | 9.79 | 351 | 0.99 |
|  | October | 47,935 | 13.22 | 567 | 1.18 |
|  | November | 28,194 | 7.78 | 474 | 1.68 |
|  | December | 5,217 | 1.44 | 182 | 3.49 |
|  | **Total** | **205,670** | **-** | **1,800** | **0.63** |
| **2022** | January | 78,135 | 21.55 | 11,967 | 15.32 |
| **Total** |  | **362,650** | **100.00** | **13,792** | **3.80** |

<sup>†</sup>by month of first episode registration; <sup>§</sup>by month of reinfection registration.

**Table 2.** **Overall registered SARS-CoV-2 primary infections (March 6, 2020-October 31,2021), SARS-CoV-2 reinfections and related hospitalization rates, in Vojvodina, Serbia, in the period March 6, 2020-January 31, 2022**.

|  |  | Overall SARS-CoV-2 primary infections <sup>†</sup> |  | Proportion of hospitalized primary infections<sup>¶</sup> |  | SARS-CoV-2 reinfections <sup>§</sup> |  | Proportion of hospitalized reinfections<sup>§</sup> |  | Primary SARS-CoV-2 infections that were reinfected |  |
| --- | --- | --- | --- | --- | --- | --- | --- | --- | --- | --- | --- |
|  | Months | n | % | n | % | n | % | n | % | n | % |
| 2020 | March-June | 1,570 | 0.63 | 1,539 | 98.02 | - | - | - | - | 80 | 5.10 |
|  | July | 4,597 | 1.83 | 2,102 | 45.73 | 0 | 0 | - | - | 296 | 6.44 |
|  | August | 1,750 | 0.70 | 420 | 24.00 | 0 | 0 | - | - | 92 | 5.26 |
|  | September | 330 | 0.13 | 66 | 20.00 | 0 | 0 | - | - | 29 | 8.79 |
|  | October | 1,547 | 0.62 | 196 | 12.67 | 1 | 0.01 | 0 | 0 | 171 | 11.05 |
|  | November | 30,659 | 12.21 | 1,796 | 5.86 | 9 | 0.07 | 1 | 11.11 | 3,272 | 10.67 |
|  | December | 38,392 | 15.29 | 2,138 | 5.57 | 15 | 0.11 | 1 | 6.67 | 3,353 | 8.73 |
|  | Total | 78,845 | - | 8,257 | 10.47 | 25 | - | 2 | 8.00 | 7,293 | 9.25 |
| 2021 | January | 11,441 | 4.56 | 868 | 7.59 | 7 | 0.05 | 0 | 0.00 | 858 | 7.50 |
|  | February | 11,868 | 4.73 | 831 | 7.00 | 11 | 0.08 | 3 | 27.27 | 925 | 7.79 |
|  | March | 30,012 | 11.95 | 2,089 | 6.96 | 47 | 0.34 | 2 | 4.25 | 1,874 | 6.24 |
|  | April | 21,929 | 8.73 | 1,691 | 7.71 | 47 | 0.34 | 2 | 4.25 | 1,211 | 5.52 |
|  | May | 5,084 | 2.02 | 491 | 9.66 | 21 | 0.15 | 0 | 0.00 | 287 | 5.65 |
|  | June | 586 | 0.23 | 70 | 11.95 | 5 | 0.04 | 1 | 20.00 | 43 | 7.34 |
|  | July | 860 | 0.34 | 71 | 8.26 | 8 | 0.06 | 0 | 0.00 | 39 | 4.53 |
|  | August | 7,029 | 2.80 | 442 | 6.28 | 80 | 0.58 | 6 | 7.50 | 187 | 2.66 |
|  | September | 35,515 | 14.14 | 2,094 | 5.90 | 351 | 2.54 | 13 | 3.70 | 657 | 1.85 |
|  | October | 47,935 | 19.09 | 3,061 | 6.39 | 567 | 4.11 | 19 | 3.35 | 418 | 0.87 |
|  | November | - | - | - | - | 474 | 3.44 | 13 | 2.74 | - | - |
|  | December | - | - | - | - | 182 | 1.32 | 6 | 3.30 | - | - |
|  | Total | 172,259 | - | 11,708 | 6.80 | 1,800 | - | 65 | 3.61 | 6,499 | 3.77 |
| 2022 | January | - | - | - | - | 11,967 | 86.77 | 82 | 0.69 | - | - |
| TOTAL |  | 251,104 | 100.00 | 19,965 | 7.95 | 13,792 | 100.00 | 149 | 1.08 | 13,792 | 5.49 |

<sup>†</sup>by month of first episode registration; <sup>§</sup>by month of reinfection registration.

The overall incidence rate of SARS-CoV-2 reinfections was 5·99 (CI 5·89-6·09) per 1,000 person-months. Fig.1A shows the Kaplan-Meier plot for the reinfection risk for patients who survived three months after primary infection (n=242,737). The risk becomes 0·76% at six months, 1·36% at nine months, 4·96% at 12 months, 16·68% at 15 months, 18·86% at 18 months. The risk of reinfection for severe or critical clinical forms of primary infections at these time points was 0·25% 0·74%, 2·26%, 6·14%, 7·10%, and 0·82%, 3·74%, 4·87%, 6·78%, 6·78%, respectively (Fig.1B). The probability of reinfection was low before, and substantially higher, after the advent of Omicron in in the sixth pandemic wave (Fig.1C).

**Figure 1.**
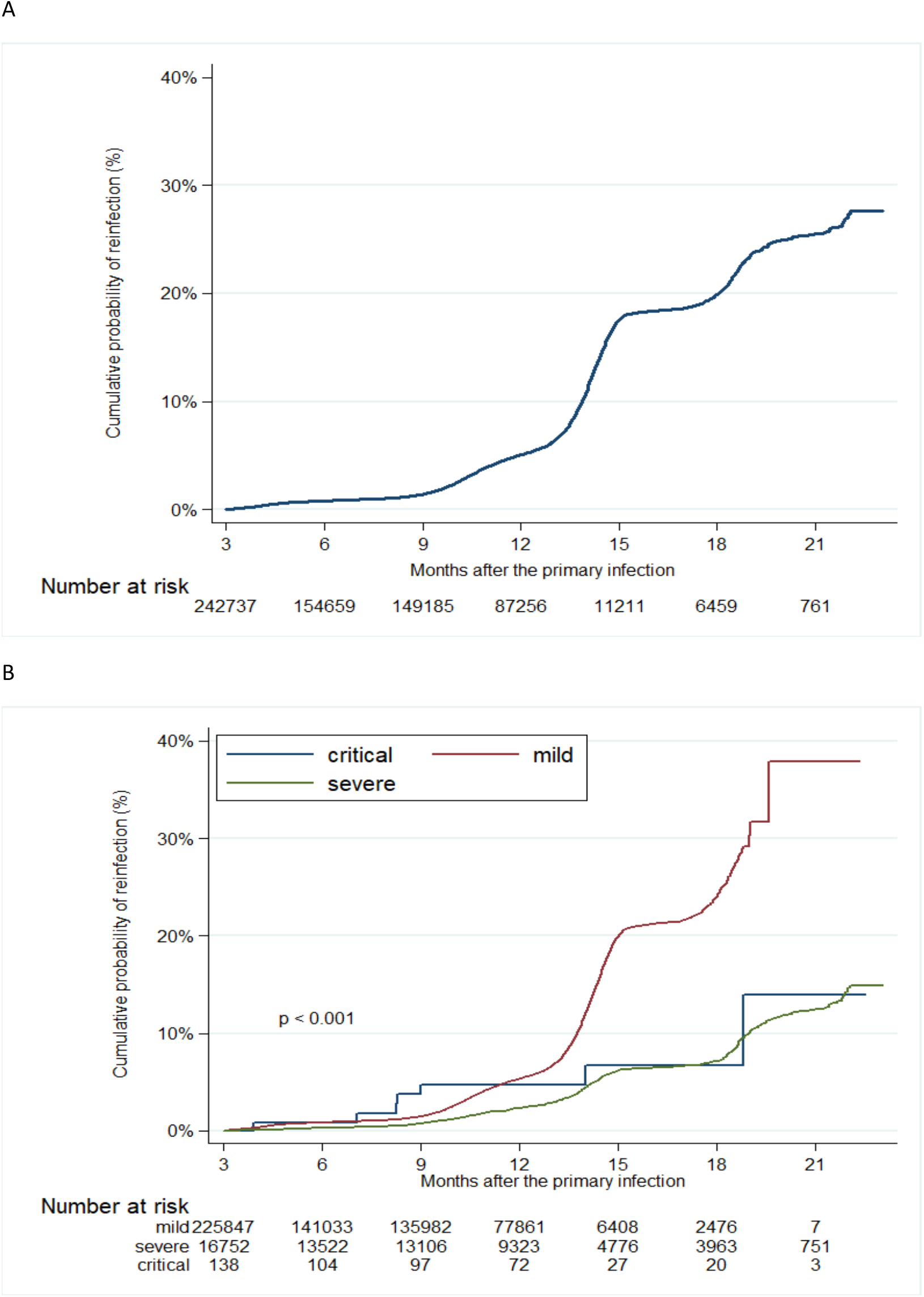
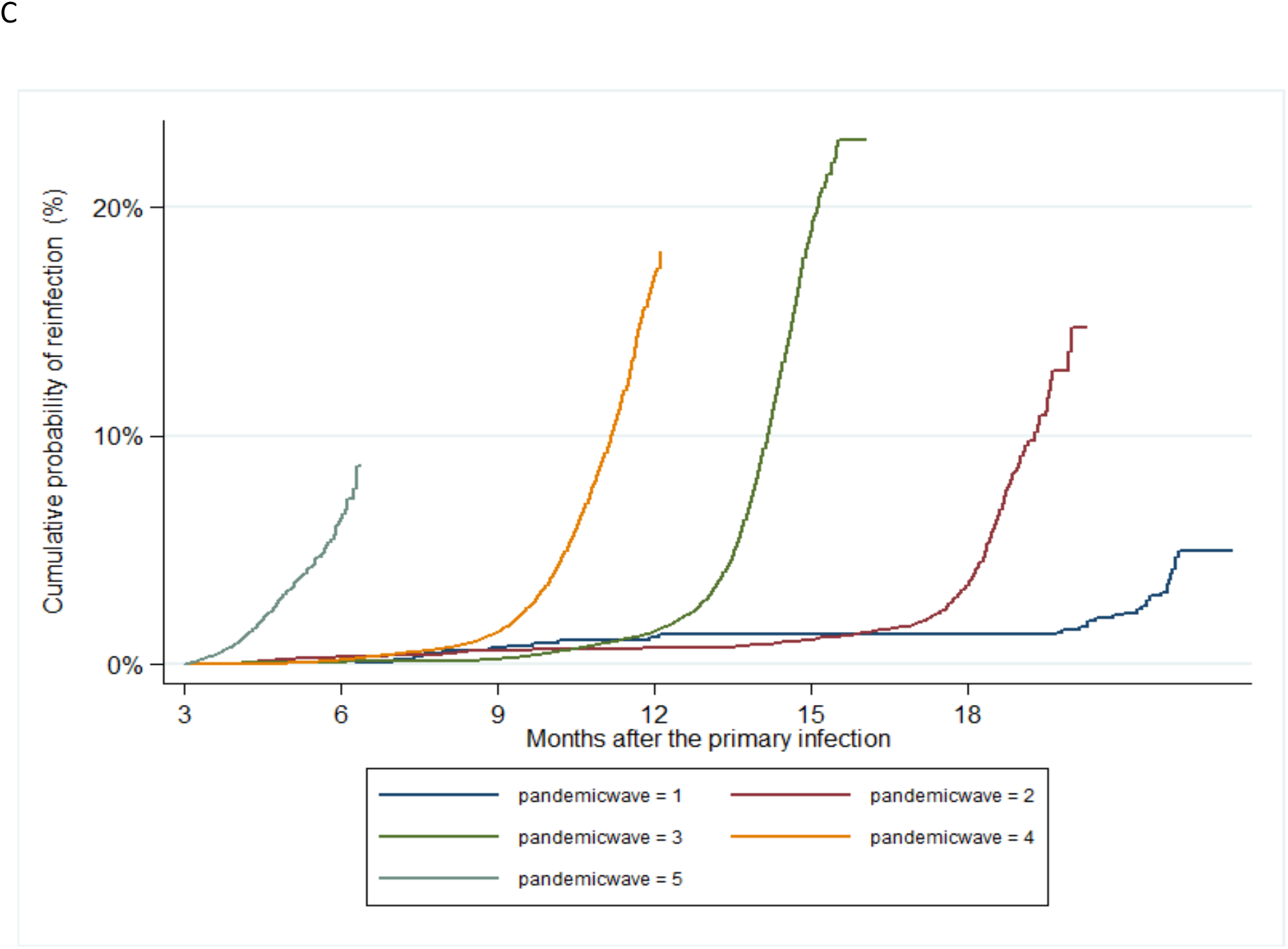
Kaplan-Meier curve showing: the cumulative probability of reinfection in the overall cohort (A) and according to disease severity (B) and pandemic waves* (C), in Vojvodina, Serbia, March 2020-January 2022. *Duration of pandemic waves: First pandemic wave lasted from March 6 until June 1, 2020; Second pandemic wave lasted from June 2 until October 6, 2020; Third pandemic wave lasted from October 7, 2020 until January 31, 2021; Forth pandemic wave lasted from February 1 until July 23, 2021; Fifth pandemic wave lasted from July 24 until December 31, 2021.

### Patients’ characteristics and risk factors for re-infection and for severe re-infection

Compared to patients without reinfection, reinfected patients were significantly younger (mean age 41·61±14·43 *vs*. 46·16 ±18·87 years), with more pronounced female dominance (57·98% *vs*. 52·65%), more frequently employed as health-care workers (HCWs) (10·90% *vs*. 4·37%), and with lower vaccination coverage (2·51% *vs*. 10·26% who received ≥2 doses) at the day of laboratory confirmation of primary infection. Other main characteristics of both groups of patients are shown in Table 3. Older age (≥70 years), ≥1 comorbidities and severe (OR 7·35, CI 4·84-11·17, p<0·001) or critical primary infection (OR 221·40, CI 48·74-1005·65, p<0·001) were significantly associated with severe reinfections (Table 4).

**Table 3.** **Demographic characteristics and vaccination status of COVID-19 cases and SARS-CoV-2 reinfections in Vojvodina, Serbia, March 2020-October 2021¶**.

| Characteristic | Overall COVID-19 cases<br>n=251,104 |  | Reinfection cases <sup>§</sup><br>n=13,792 |  | Cases without reinfection <sup>§</sup><br>n=237,312 |  | P value |
| --- | --- | --- | --- | --- | --- | --- | --- |
|  | n | % | n | % | n | % |  |
| **Gender** |  |  |  |  |  |  |  |
| Male | 118,152 | 47.05 | 5,795 | 42.02 | 112,357 | 47.35 | <0.001 |
| Female | 132,952 | 52.95 | 7,997 | 57.98 | 124,955 | 52.65 |  |
| **Age (years), mean ± SD** | 45.91± 18.68 |  | 41.61±14.43 |  | 46.16±18.87 |  | <0.001 |
| **Age category** | n | % | n | % | n | % |  |
| 0-9 | 4,220 | 1.68 | 105 | 0.76 | 4,115 | 1.73 | <0.001 |
| 10-19 | 17,633 | 7.02 | 661 | 4.79 | 16,972 | 7.15 |  |
| 20-29 | 29,490 | 11.74 | 1,976 | 14.33 | 27,514 | 11.59 |  |
| 30-39 | 45,235 | 18.01 | 3,693 | 26.78 | 41,542 | 17.51 |  |
| 40-49 | 48,262 | 19.22 | 3,516 | 25.49 | 44,746 | 18.86 |  |
| 50-59 | 40,687 | 16.20 | 2,189 | 15.87 | 38,498 | 16.22 |  |
| 60-69 | 36,702 | 14.62 | 1,189 | 8.62 | 35,513 | 14.96 |  |
| 70-79 | 20,450 | 8.14 | 361 | 2.62 | 20,089 | 8.47 |  |
| ≥80 | 8,425 | 3.36 | 102 | 0.74 | 8,323 | 3.51 |  |
| **Occupation** |  |  |  |  |  |  |  |
| Healthcare workers (HCWs) | 11,872 | 4.73 | 1,504 | 10.9 | 10,368 | 4.37 | <0.001 |
| Non-HCWs<sup>#</sup> | 187,924 | 74.84 | 11,215 | 81.32 | 176,709 | 74.46 |  |
| Retired | 51,308 | 20.43 | 1,073 | 7.78 | 50,235 | 21.17 |  |
| **Comorbidities** |  |  |  |  |  |  |  |
| 0 | 198,017 | 78.86 | 11,514 | 83.48 | 186,503 | 78.59 | <0.001 |
| 1 | 39,828 | 15.86 | 1,803 | 13.07 | 38,025 | 16.02 |  |
| 2 | 10,123 | 4.03 | 372 | 2.7 | 9,751 | 4.11 |  |
| ≥3 | 3,136 | 1.25 | 103 | 0.75 | 3,033 | 1.28 |  |
| **Vaccination status** |  |  |  |  |  |  |  |
| Unvaccinated | 213,268 | 84.93 | 13,207 | 95.76 | 200,061 | 84.30 | <0.001 |
| Partially vaccinated | 13,154 | 5.24 | 238 | 1.73 | 12,916 | 5.44 |  |
| Fully vaccinated | 21,846 | 8.7 | 325 | 2.35 | 21,521 | 9.10 |  |
| Boosted | 2,836 | 1.13 | 22 | 0.16 | 2,814 | 1.16 |  |

<sup>¶</sup> Indicators of significance between groups using Pearson's chi-squared test and unpaired t-test.
<sup>§</sup> At the time of laboratory confirmation of primary infection
<sup>#</sup>Non-HCWs included professions that provide various services, other employment categories as well as retired individuals.

**Table 4.** **Associations between different factors and the severity of the reinfection (i.e. severe+critical *vs*. mild) in Vojvodina, Serbia, Mar 2020-Jan 2022**.

|  | Unadjusted Model |  |  |  | Adjusted Model<sup>§</sup> |  |  |  |
| --- | --- | --- | --- | --- | --- | --- | --- | --- |
|  | OR | 95% CI |  | p-value | OR | 95% CI |  | p-value |
| Gender |  |  |  |  |  |  |  |  |
| Male | 1.43 | 0.99 | 2.07 | 0.056 | - | - | - | - |
| Female | Reference group |  |  |  | - | - | - | - |
| Age (years) | 1.06 | 1.04 | 1.07 | <0.001 | - | - | - | - |
| Age category (years) |  |  |  |  |  |  |  |  |
| 0-9 | NA | NA | NA | NA | - | - | - | - |
| 10-19 | Reference group |  |  |  | - | - | - | - |
| 20-29 | 4.03 | 0.52 | 31.07 | 0.181 | - | - | - | - |
| 30-39 | 3.23 | 0.43 | 24.26 | 0.254 | - | - | - | - |
| 40-49 | 2.83 | 0.37 | 21.44 | 0.315 | - | - | - | - |
| 50-59 | 4.86 | 0.64 | 36.71 | 0.125 | - | - | - | - |
| 60-69 | 14.75 | 2.00 | 108.98 | 0.008 | - | - | - | - |
| 70-79 | 38.71 | 5.17 | 289.66 | <0.001 | - | - | - | - |
| ≥80 | 41.25 | 4.91 | 346.35 | 0.001 | - | - | - | - |
| Occupation |  |  |  |  |  |  |  |  |
| Health-care workers (HCWs) | 0.81 | 0.37 | 1.78 | 0.608 | 0.87 | 0.40 | 1.93 | 0.737 |
| Non-HCWs<sup>#</sup> | Reference group |  |  |  | Reference group |  |  |  |
| Retired | 7.27 | 4.92 | 10.76 | <0.001 | 3.00 | 1.63 | 5.52 | <0.001 |
| Number of comorbidities |  |  |  |  |  |  |  |  |
| 0 | Reference group |  |  |  | Reference group |  |  |  |
| 1 | 2.81 | 1.82 | 4.32 | <0.001 | 1.58 | 1.00 | 2.50 | 0.051 |
| 2 | 5.53 | 2.97 | 10.30 | <0.001 | 2.59 | 1.35 | 4.99 | 0.004 |
| ≥3 | 4.98 | 1.54 | 16.07 | 0.007 | 2.10 | 0.64 | 6.93 | 0.224 |
| Severity of primary infection |  |  |  |  |  |  |  |  |
| Mild | Reference group |  |  |  | Reference group |  |  |  |
| Severe | 7.35 | 4.84 | 11.17 | <0.001 | 5.01 | 3.26 | 7.71 | <0.001 |
| Critical | 221.40 | 48.74 | 1005.65 | <0.001 | 189.00 | 39.13 | 912.95 | <0.001 |

<sup>§</sup>adjusted for age (continuous) and gender.

### Severity of reinfections versus primary infections

Most reinfections were mild (99·17%), and only a minority were severe (0·78%) or critical (0·05%). Reinfections were milder than primary infections. The share of severe clinical forms decreased from 5·47% in initial episodes to 0·78% in reinfections, while critical forms remained the same (0·05%). The proportion of severe and critical disease forms was higher in cases without reinfection (9·30 and 0·70%, respectively) (Fig.2).

**Figure 2.**
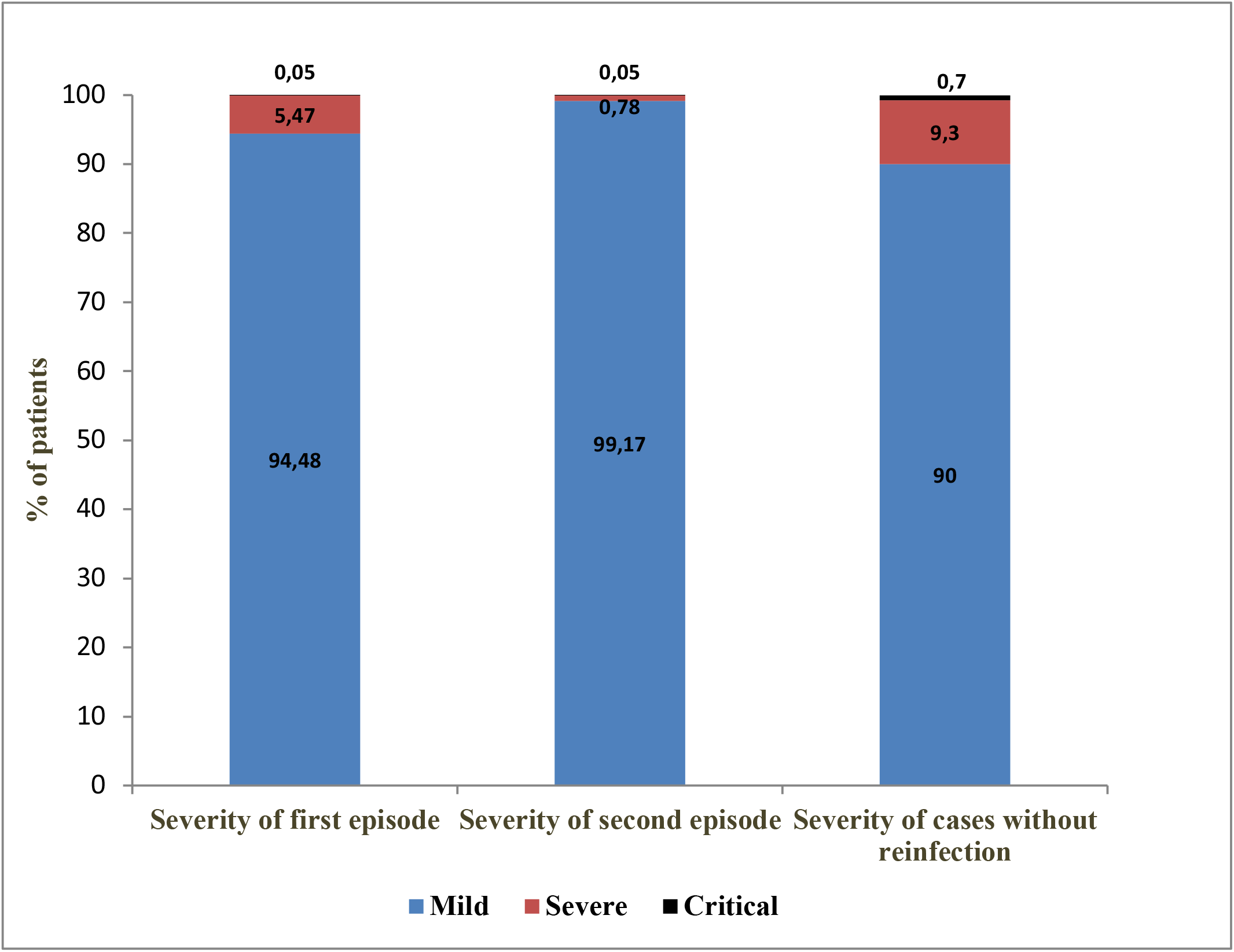
Severity of COVID-19 in patients with reinfection (first and second episode) and patients without reinfection, in Vojvodina, Serbia, March 06, 2020-January 31, 2022.

### Hospitalizations after reinfection

Overall, the hospitalization rate of COVID-19 patients in the observed period was 7·95%. At the beginning of the pandemic (March-June 2020) almost all patients were hospitalized (98·02%). The proportion of hospitalized gradually decreased from 10·47% in 2020 to 6·80% in 2021 (Table 2). Reinfected patients were rarely hospitalized (1·08% *vs*. 3·70% during initial infection). Reinfected patients were 4·2 times more likely to be hospitalized during initial infection compared to reinfection (McNemar OR=4·21, 95%CI 3·41-5·22, p<0.001). Overall, 38 patients were hospitalized in both the primary infection and reinfection. In January 2022, when most reinfections were recorded, only 0·69% of patients were hospitalized.

### Deaths

Among reinfected patients, 31 deaths occurred, of which 20 were classified as COVID-19 deaths (case fatality ratio 0·15% among 13,792 reinfections); the rest were caused by non-COVID causes. COVID-19 deaths occurred mainly in reinfected patients ≥60 years (70·0%), with severe (75·0%) or critical disease (25·0%). Most of them died during the fifth (50·00%) or sixth (35·00%) pandemic wave (SI Table 1). K-M curves for the risk of death for the entire cohort of 251,104 positive patients and for reinfected patients appear in SI Fig.2.

### Second reinfections

In total, 34 cases (0·01%) with three consecutive SARS-CoV-2 reinfections were recorded (SI Table 2). Cases of second reinfection ≥90 days after the previous one were registered in December 2021 (n=1) and January 2022 (n=33). Overall, 14·7% of patients were hospitalized due to severe primary infection, while first and second reinfections were mild. None of these reinfections led to hospitalization or death. On the day of laboratory confirmation of the second reinfection, most patients (55·9%) were unvaccinated.

### Case control study

A total of 13,189 reinfections were recorded in the period January 1, 2021, through January 31, 2022, of which 7071 (53·6%) met the eligibility criteria for study cases and were matched with 14,142 controls. Women (56·8%) and middle-age groups (55·0%) predominated in the study population, while most participants (91·4%) were initially infected during October–December 2020 (SI Table 3). In relation to vaccination status of study cases and controls, the distribution of unvaccinated (≈52%) and incompletely vaccinated (≈2%) was similar but there were more fully vaccinated (24·7% *vs*. 20·3%), and less boosted (21·3% *vs*. 25·9%) (p<0.001) among study cases. All three categories of cases, regardless of whether they were unvaccinated (OR=1·23; 95%CI=1·14–1·33), incompletely (OR=1·33; 95% CI=1·08–1·64) or completely vaccinated (OR=1·50; 95% CI=1·37–1·63), were modestly more likely to be reinfected compared with those who received a booster dose (SI Table 4).

## Discussion

In this large population-based study, we have documented that the risk of reinfection remained exceedingly low before the emergence of Omicron and increased substantially thereafter, accounting for 15% of the infections during January 2022. Reinfections were generally mild, and their severity was much lower compared with primary infections. Accordingly, hospitalizations were uncommon and fatal outcomes were distinctly rare, much lower than in primary infections. Vaccination, in particular a booster dose, diminished modestly the probability of reinfection in a case-control analysis.

In the early stages of the pandemic, accumulated evidence supported the effectiveness of natural immunity after infection in protecting against reinfection with different SARS-CoV-2 variants for at least one year.^3^ Omicron, however, showed substantial ability to evade natural and vaccine-induced immunity, leading to reduced protection against reinfection, but similar protection against hospitalization or death due to reinfection.^18^ Unlike the Beta and Delta periods of predominance when there was no evidence of population-level immune escape, the explosive spread of Omicron in South Africa led to a dramatic increase of reinfections in mid-November 2021.^19^ Similar findings have also been reported in Qatar and the United Kingdom and are relevant to the dominance of Omicron worldwide by early 2022.^18,20^

Repeated infections should be anticipated from viruses like SARS-CoV-2 that infect mucosal surfaces without a viremic phase, which typically result in relatively short-lived antibody responses.^21^ Accordingly, we registered 34 patients with three consecutive infections, almost all in January 2022. An increased risk of a third infection was also documented in South Africa, with 1,788 individuals with at least three and 18 with four suspected infections.^19^ Almost all third infections occurred after October 2021, during the period of Omicron circulation.

The global epidemiology of SARS-CoV-2 in January 2022 was characterized by the emergence of Omicron (B.1.1.529), declining prevalence of Delta (B.1.617.2), and very low-level circulation of previous VOCs.^22^ The only published genomic surveillance data from Serbia thus far revealed the existence of different Alpha sub-lineages (B.1.1.1 and B.1.1.7) in the period April-October 2020.^23,24^ Despite the limited availability of sequencing data from Serbia, it seems reasonable to assume that the increase in reinfections after August 2021 was due to introduction and spread of Delta, while the abrupt increase in January 2022 was due to the explosive spread of Omicron.

Characteristics that we observed to be more common among reinfected patients (younger, working-age groups, more often employed as HCWs) support the hypothesis that the reinfection risk is a function of the risk of exposure.^3^ Older age groups, especially retirees, were less exposed due to lockdowns in 2020 and generally more compliant with the recommendations of mask wearing, distancing, and accepting vaccination. In our study, being older ≥70 years, having ≥1 comorbidities and a severe or critical primary infection were significantly associated with severe reinfections. In a retrospective cohort study of SARS-CoV-2-positive HCWs in 2020, reinfections were uncommon and more likely in women, adults, immunocompromised and previously hospitalized for COVID-19.^25^ More frequent testing may also affect the documentation of reinfections. Most reinfections are likely to be missed otherwise, if they are asymptomatic or have very mild symptoms.

Clinical manifestations of SARS-CoV-2 reinfections compared to primary infections vary from mild to severe and even life-threatening, yet factors contributing to changes in severity remain largely unknown. In our study, reinfections tended to be milder compared to primary infections. Similarly, reinfections had 90% lower odds of resulting in hospitalization or death compared to primary infections in Qatar and were generally rare and mild.^26^ A plausible explanation for this finding could be a primed immune system after the initial infection, which raises hope that the disease course could be milder when the virus becomes endemic. The massive surges of Omicron in late 2021 and 2022 that were often accompanied by very mild disease and disproportionately few severe cases may be consistent with a transition to endemicity. Perhaps as exceptions to the general rule, severe reinfections in different age and racial/ethnic groups have also been reported.^27,28^

A recent systematic review showed wide variations between the severity of primary infection compared to reinfection, with increases in asymptomatic cases.^29^ In a study by Wang et al., the majority (69%) of reinfections with a genetically distinct SARS-CoV-2 strain, were of similar severity; 18·8% had worse and 12·5% milder symptoms with the second episode.^30^ Additionally, in a pre-print of a cohort study on the immune correlates of natural infection, seropositivity was associated with 69·2% protection from symptomatic infection mostly against Gamma and Delta variants with higher protection against moderate or severe infection and reinfections were less severe than first infections.^31^

The frequency and timeliness of required vaccination doses (typically for protection against severe COVID-19) have not yet been determined with certainty. The third dose should probably be seen as the final dose of the original vaccination scheme. We observed a slightly decreased reinfection risk in boosted versus unvaccinated, partially and fully vaccinated people. This result, however, needs to be interpreted with caution since individuals may vary in other factors that were not accounted for in our analysis.^32^ For example, boosted people may be more health conscious in general and have a heightened sense of protection, resulting in lower reinfection rates. Several studies have shown vaccinated people to be in better general health, e.g. having three-fold lower mortality risk from non-COVID-19 causes than non-vaccinated.^33^ Still, our data are compatible with the potential benefits of hybrid immunity, generated from the combination of prior infection plus vaccination. If true, the modest benefit of a booster dose on a relative risk scale needs to be examined also in terms of absolute risk reduction, a task that might not be trivial currently, in the absence of an active epidemic wave in most countries. Randomized trials should be considered to evaluate booster doses in the future.

We must emphasize some limitations. Both primary infections and reinfections were diagnosed mainly by Ag-RDT testing without subsequent confirmative RT-PCR testing of positive results. There is a lack of evidence of genotyping variance, threshold cycle values as well as at least one negative between two positive RT-PCR tests in patients with suspected reinfection.^34^ Therefore, the possibility that reinfection was caused by genetically different SARS-CoV-2 variants compared to primary infection could not be investigated. Most asymptomatic patients and those who did not seek testing were not captured, particularly in the early pandemic days. Also, if previously infected people were tested less due to their presumed natural immunity, the reinfection rate could have been underestimated. Given missed asymptomatic reinfections, the proportion of severe reinfections is certainly substantially over-estimated many-fold. Matching was not implemented to control for differences in race, nationality and education level that might influence the decision to be vaccinated or vaccine choice.

Our study offers large-scale population-level evidence on reinfections over a two-year period. Since spatiotemporal differences are relevant to SARS-CoV-2 reinfections, longer prospective population-based studies with well characterized virologic and immunologic data are needed to assess the risk of reinfection in the future and whether low severity remains a key feature.

## Data Availability

All data produced in the present study are available upon reasonable request to the authors.

## Data sharing statement

The data that support the findings of this study are available upon request with approval needed from the Center for Disease Control and Prevention, Institute of Public Health of Vojvodina. The data are not publicly available due to restrictions pertaining to contained information that could compromise the privacy of patients.

## Author contributions

SM, CA and JPAI conceived and designed the study. SM, VV, NG, VP, TP, MR, TP and ZG processed laboratory results and collected data, SM, CA, ZLC, ND, VV, AT analyzed and interpreted data, SM and CA supervised and validated data. SM, CA, ZLC, ND, VV, TP and JPAI were involved in data visualization, presentation and formal analysis. SM, ZLC, VV, ND and VP accessed and verified underlying data. SM, CA, AT and JPAI drafted the Article. All authors critically reviewed the article. All authors had access to aggregated data reported in the study and had final responsibility for the decision to submit the publication.

## Acknowledgments

The authors would like to thank all epidemiologists and health-care workers who were involved in the study. We also wish to thank computer engineer Dušan Krstić for the excellent technical assistance. The work of Zagorka Lozanov-Crvenković was funded by the Ministry of Education, Science and Technological Development of the Republic of Serbia (grant numbers: 451-03-68/2022-14/200032 and 451-03-68/2022-14/200125).

## Supplementary material

**Supplementary Table 1.**
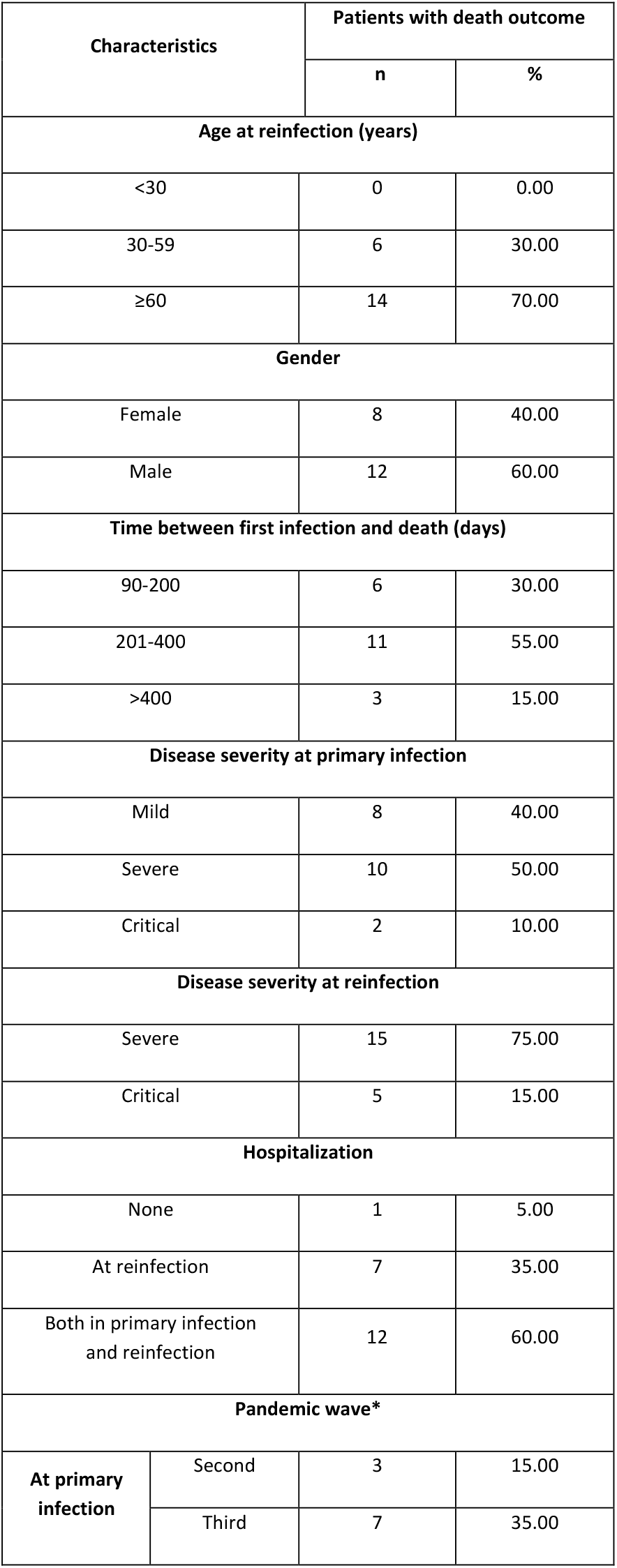
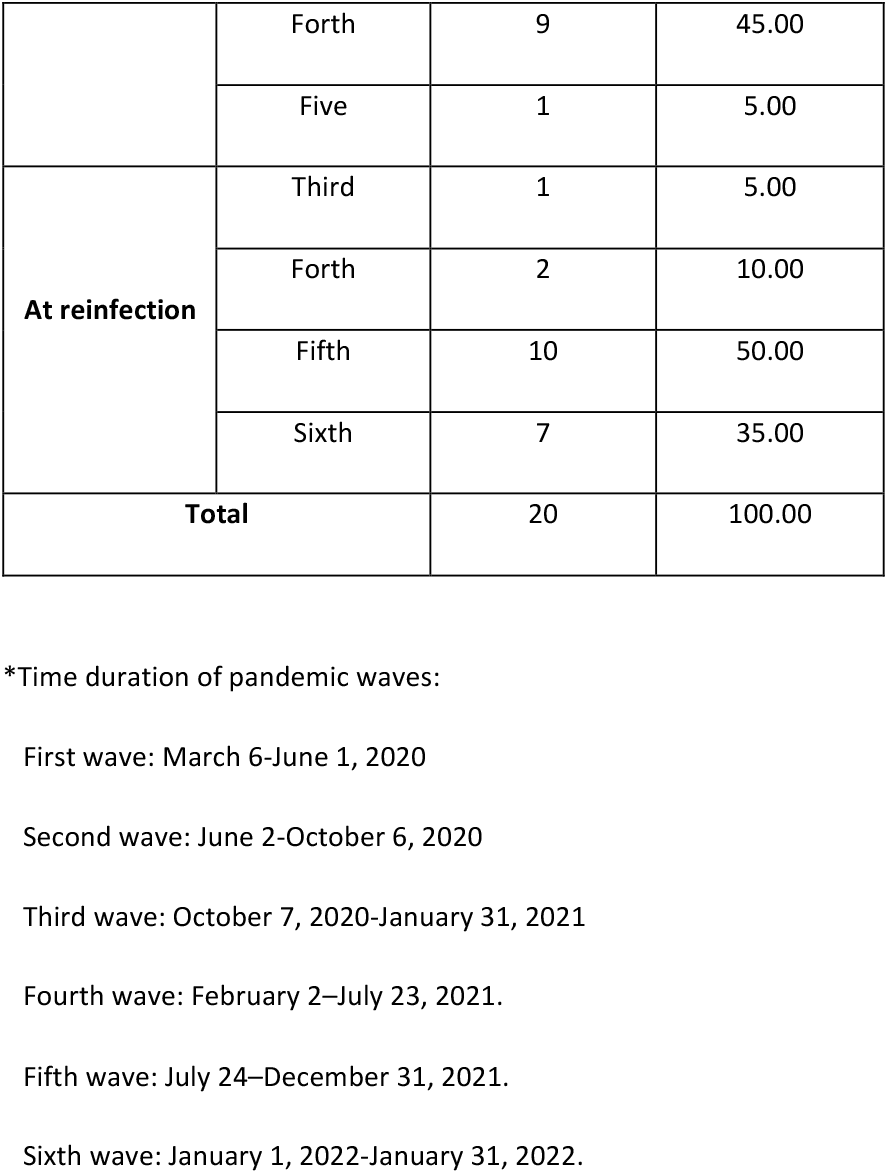
**Characteristics of patients with death outcome caused by SARS CoV-2 reinfections, in Vojvodina, Serbia, March 6, 2020 - January 31,2022**.

| Characteristics |  | Patients with death outcome |  |
| --- | --- | --- | --- |
|  |  | n | % |
| **Age at reinfection (years)** |  |  |  |
| <30 |  | 0 | 0.00 |
| 30-59 |  | 6 | 30.00 |
| ≥60 |  | 14 | 70.00 |
| **Gender** |  |  |  |
| Female |  | 8 | 40.00 |
| Male |  | 12 | 60.00 |
| **Time between first infection and death (days)** |  |  |  |
| 90-200 |  | 6 | 30.00 |
| 201-400 |  | 11 | 55.00 |
| >400 |  | 3 | 15.00 |
| **Disease severity at primary infection** |  |  |  |
| Mild |  | 8 | 40.00 |
| Severe |  | 10 | 50.00 |
| Critical |  | 2 | 10.00 |
| **Disease severity at reinfection** |  |  |  |
| Severe |  | 15 | 75.00 |
| Critical |  | 5 | 15.00 |
| **Hospitalization** |  |  |  |
| None |  | 1 | 5.00 |
| At reinfection |  | 7 | 35.00 |
| Both in primary infection and reinfection |  | 12 | 60.00 |
| **Pandemic wave\*** |  |  |  |
| **At primary infection** | Second | 3 | 15.00 |
|  | Third | 7 | 35.00 |
|  | Forth | 9 | 45.00 |
|  | Five | 1 | 5.00 |
| **At reinfection** | Third | 1 | 5.00 |
|  | Forth | 2 | 10.00 |
|  | Fifth | 10 | 50.00 |
|  | Sixth | 7 | 35.00 |
| **Total** |  | 20 | 100.00 |

\*Time duration of pandemic waves:
First wave: March 6-June 1, 2020
Second wave: June 2-October 6, 2020
Third wave: October 7, 2020-January 31, 2021
Fourth wave: February 2-July 23, 2021.
Fifth wave: July 24-December 31, 2021.
Sixth wave: January 1, 2022-January 31, 2022.

**Supplementary Table 2.** **Characteristics of patients with three consecutive SARS-CoV-2 infections in Vojvodina, Serbia, March 6, 2020-January 31, 2022**.

| Characteristics | Thrice infected with SARS-CoV-2 |  |
| --- | --- | --- |
|  | n | % |
| **Age (years)** |  |  |
| 0-9 | 0 | 0.0 |
| 10-19 | 1 | 2.9 |
| 20-29 | 6 | 17.7 |
| 30-39 | 8 | 23.5 |
| 40-49 | 8 | 23.5 |
| 50-59 | 8 | 23.5 |
| ≥60 | 3 | 8.9 |
| **Gender** |  |  |
| Male | 11 | 32.3 |
| Female | 23 | 67.7 |
| **Severity** |  |  |
| Mild | 34 | 100.0 |
| **Profession** |  |  |
| HCWs<sup>a</sup> | 9 | 26.5 |
| Non-HCWs<sup>b</sup> | 25 | 73.5 |
| **Number of comorbidities** |  |  |
| 0 | 33 | 97.1 |
| 1 | 1 | 2.9 |
| **Vaccination status** |  |  |
| Unvaccinated | 19 | 55.9 |
| Partially vaccinated | 8 | 23.5 |
| Fully vaccinated | 4 | 11.8 |
| Boosted | 3 | 8.8 |
| **Total** | **34** | **100.0** |

<sup>a</sup> Health-care-workers (HCWs); <sup>b</sup> other occupations except HCWs.

**Supplementary Table 3.** **Characteristics of study cases (reinfected with SARS-CoV-2) and control participants (who were not reinfected) in Vojvodina, Serbia, January 1, 2021–January 31, 2022**.

| Characteristic | Study cases<sup>a</sup><br>(n=7,071) |  | Controls<br>(n=14,142) |  |
| --- | --- | --- | --- | --- |
|  | n | % | n | % |
| **Age group (years)** |  |  |  |  |
| 18-29 | 1,108 | 15.7 | 2,293 | 16.2 |
| 30-39 | 2,036 | 28.8 | 3,873 | 27.4 |
| 40-49 | 1,855 | 26.2 | 3,712 | 26.2 |
| 50-59 | 1,259 | 17.8 | 2,552 | 18.0 |
| 60-69 | 586 | 8.3 | 1,261 | 8.9 |
| 70-79 | 179 | 2.5 | 354 | 2.5 |
| ≥80 | 48 | 0.7 | 97 | 0.7 |
| **Gender** |  |  |  |  |
| Male | 3,057 | 43.2 | 6,114 | 43.2 |
| Female | 4,014 | 56.8 | 8,028 | 56.8 |
| **Occupation** |  |  |  |  |
| Healthcare workers (HCWs) | 1,021 | 14.4 | 1,510 | 10.7 |
| Non-HCWs<sup>b</sup> | 6,050 | 85.6 | 12,632 | 89.3 |
| **Number of comorbidities** |  |  |  |  |
| 0 | 5,715 | 80.8 | 10,951 | 77.4 |
| 1 | 1,071 | 15.1 | 2,476 | 17.5 |
| 2 | 225 | 3.2 | 555 | 3.9 |
| ≥3 | 60 | 0.8 | 160 | 1.1 |
| **Month of initial infection in 2020** |  |  |  |  |
| March | 0 | 0.0 | 0 | 0.0 |
| April | 13 | 0.2 | 26 | 0.2 |
| May | 2 | 0.0 | 1 | 0.0 |
| June | 44 | 0.6 | 93 | 0.7 |
| July | 268 | 3.8 | 534 | 3.8 |
| August | 86 | 1.2 | 173 | 1.2 |
| September | 23 | 0.3 | 41 | 0.3 |
| October | 170 | 2.4 | 352 | 2.5 |
| November | 3,198 | 45.2 | 6852 | 48.5 |
| December | 3,267 | 46.2 | 6070 | 42.9 |

<sup>a</sup> Each individual study case was matched to a control appropriate for age ( $\pm 3$ years), gender, and date of testing ( $\pm 10$ days).
<sup>b</sup> Non-HCWs included professions that provide various services (e.g. caregivers, waiters), other employment categories as well as retired individuals.

**Supplementary Table 4.**
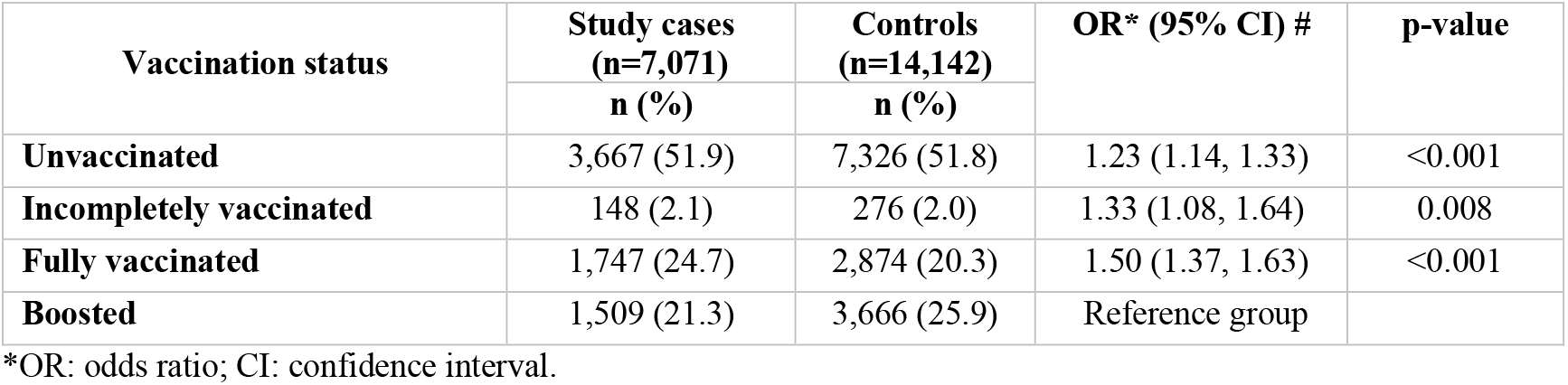
**Association of SARS-CoV-2 reinfection with COVID-19 vaccination status of study cases and controls, Vojvodina, Serbia, January 1, 2021–January 31, 2022**.

**Supplementary Figure 1:**
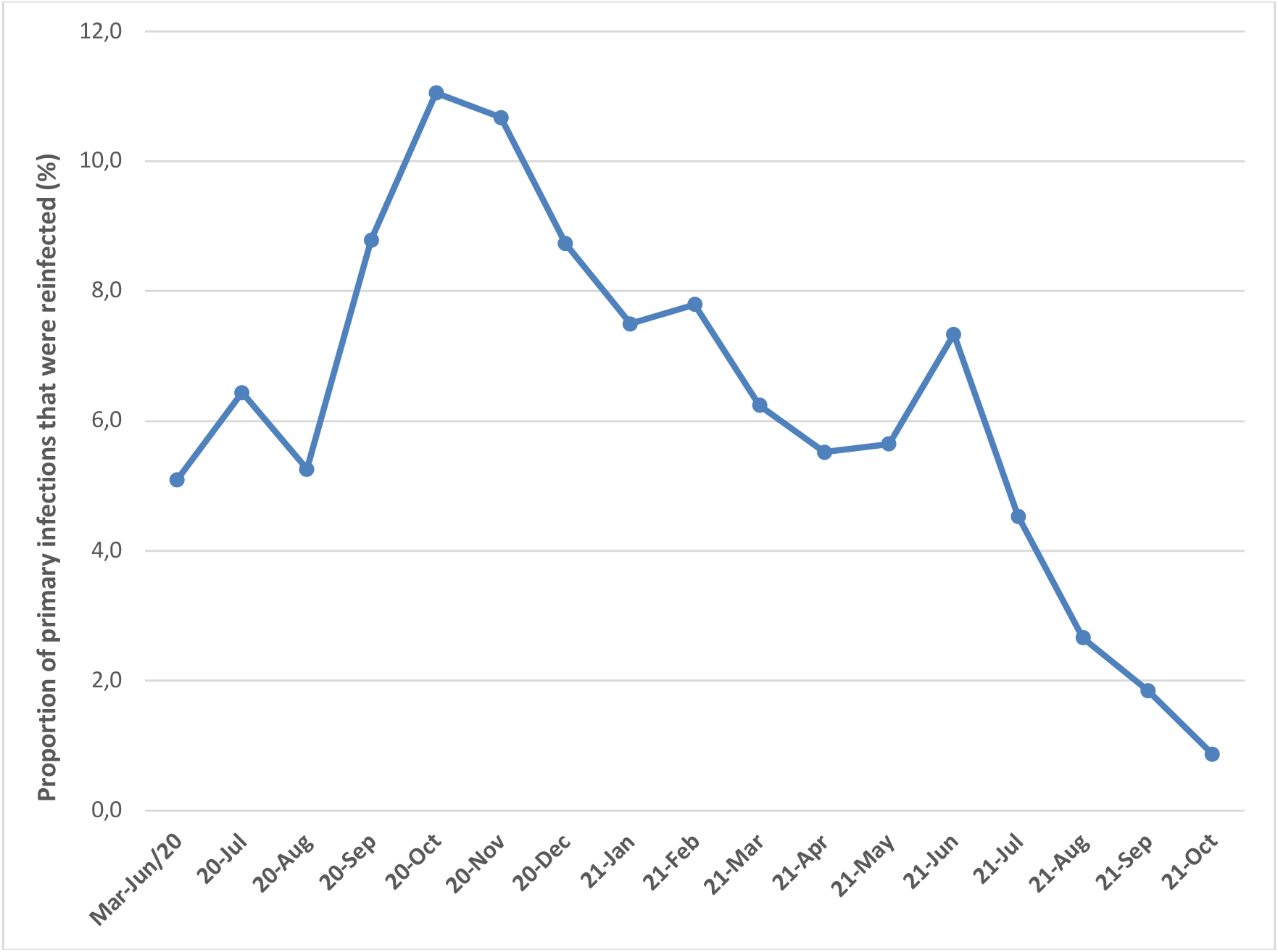
The proportion of primary infections that were reinfected in Vojvodina, Serbia, March 6, 2020-October 31, 2021.^*^ ^*^Cochran–Armitage test was applied

**Supplementary Figure 2.**
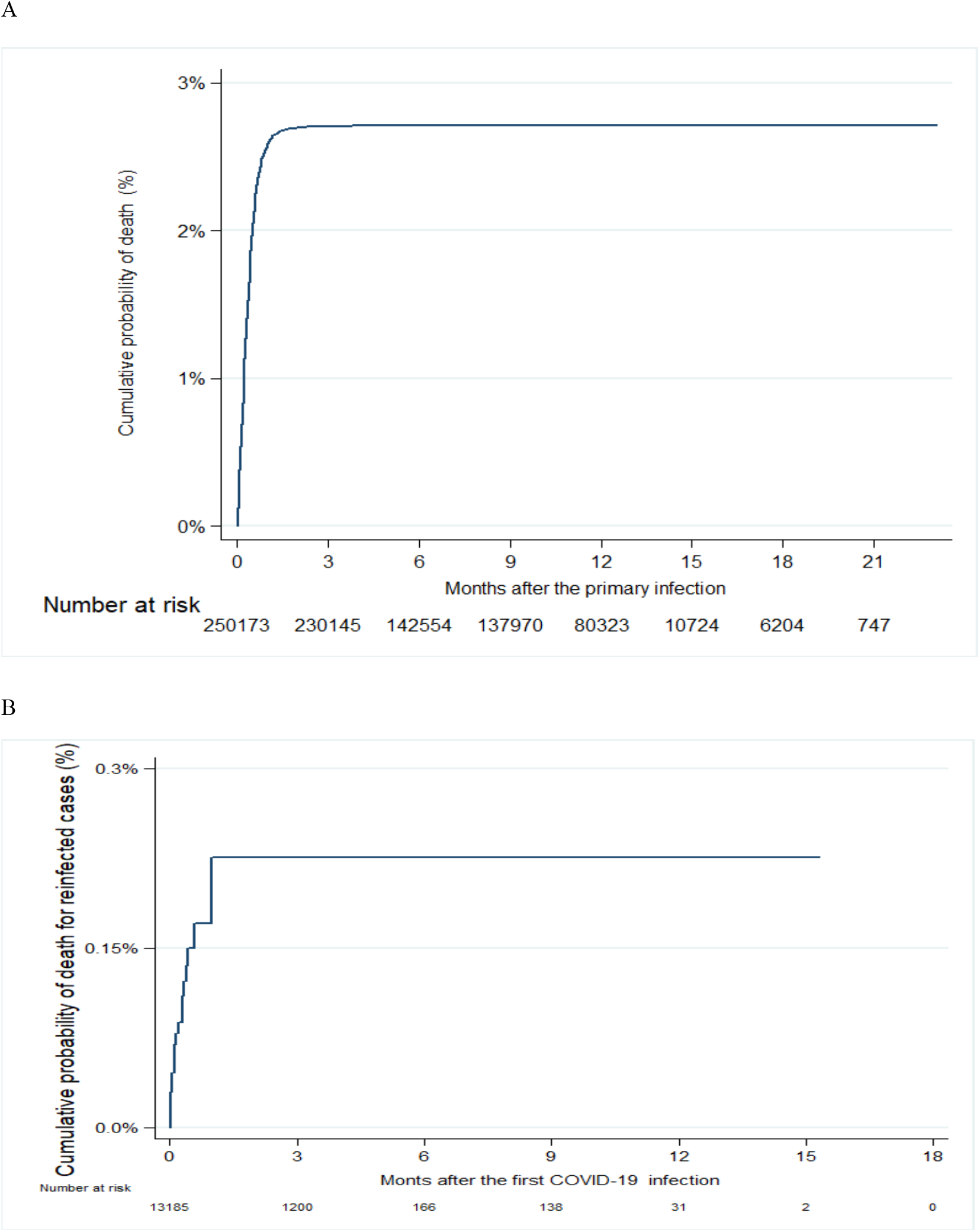
Kaplan-Meier curve showing: the cumulative probability of death in the overall cohort (A) and for the patients with reinfection (B) in Vojvodina, Serbia, March 2020-January 2022.

## References

1. European Centre for Disease Prevention and Control E. Reinfection with SARSCoV-2: considerations for public health response 2020. Available at: https://www.ecdc.europa.eu/sites/default/files/documents/Re-infection-and-viral-shedding-threat-assessment-brief.pdf (accessed March 12, 2022).

2. Yahav D, Yelin D, Eckerle I, et al. Definitions for coronavirus disease 2019 reinfection, relapse and PCR re-positivity. Clin Microbiol Infect 2021; 27(3): 315–318.

3. Pilz S, Theiler-Schwetz V, Trummer C, Krause R, Ioannidis JPA. SARS-CoV-2 reinfections: Overview of efficacy and duration of natural and hybrid immunity. Environ Res 2022; 209: 112911.

4. Krammer F. Correlates of protection from SARS-CoV-2 infection. Lancet 2021; 397: 1421–1423.

5. O Murchu E, Byrne P, Carty PG, et al. Quantifying the risk of SARS-CoV-2 reinfection over time. Rev Med Virol 2022; 32(1): e2260.

6. Pilz S, Chakeri A, Ioannidis JP, et al. SARS-CoV-2 re-infection risk in Austria. Eur J Clin Invest 2021; 51 (4): e13520.

7. Hansen CH, Michlmayr D, Gubbels SM, Mølbak K, Ethelberg S. Assessment of protection against reinfection with SARS-CoV-2 among 4 million PCR-tested individuals in Denmark in 2020: a population-level observational study. Lancet 2021; 397 (10280): 1204–1212.

8. Cavanaugh AM, Spicer KB, Thoroughman D, Glick C, Winter K. Reduced Risk of Reinfection with SARS-CoV-2 After COVID-19 Vaccination - Kentucky, May-June 2021. MMWR Morb Mortal Wkly Rep 2021; 70 (32): 1081–1083.

9. Ronchini C, Gandini S, Pasqualato S, et al. Lower probability and shorter duration of infections after COVID-19 vaccine correlate with anti-SARS-CoV-2 circulating IgGs. PLoS One 2022; 17 (1): e0263014.

10. Bio-Tech Corp, Ltd. Liferiver Novel Coronavirus (2019-nCoV) Real Time Multiplex RT-PCR Kit. Shanghai: Bio-Tech Corp, Ltd; 2020 https://www.who.int/diagnostics_laboratory/eual/eul_0486_139_00_novel_coronavirus_sars_cov_2_real_time_multiplex_rt_pcr_kit_ifu.pdf (accessed December 30, 2021).

11. OSANG Healthcare Corp, Ltd. GeneFinder COVID-19 Plus RealAmp Kit - Instructions for Use. Anyang-si: OSANG Healthcare Corp, Ltd; 2020 https://www.fda.gov/media/137116/download. BioMérieux, Inc. ARGENE® SARS-COV-2 R-GENE (accessed December 31, 2021].

12. Durham, North Carolina: BioMérieux, Inc; 2020. https://www.fda.gov/media/137742/download (accessed December 29, 2021).

13. BGI Genomics Corp Ltd. Real-Time Fluorescent RT-PCR Kit for Detecting SARS-CoV-2 - Instructions for Use. Shenzhen: BGI Genomics Corp Ltd; 2021 https://www.fda.gov/media/136472/download (accessed December 29, 2021).

14. Ristić M, Nikolić N, Ćabarkapa V, Turkulov V, Petrović V. Validation of the STANDARD Q COVID-21 antigen test in Vojvodina, Serbia. PLoS One 2021; 16 (2): e0247606.

15. Pustahija T, Ristić M, Medić S, et al. Epidemiological characteristics of COVID-19 travel-associated cases in Vojvodina, Serbia, during 2020. PLoS One 2021; 16 (12): e0261840.

16. World Health Organization. Antigen-detection in the diagnosis of SARS-CoV-2 infection using rapid immunoassays. Interim guidance. 11 September 2020. Geneva; 2020. https://www.who.int/publications/i/item/antigen-detection-in-the-diagnosis-of-sars-cov-2infection-using-rapid-immunoassays (accessed February 11, 2022).

17. Benjamin DJ, Berger JO, Johannesson M, et al. Redefine statistical significance. Nat Hum Behav 2018; 2 (1): 6–10.

18. Altarawneh HN, Chemaitelly H, Hasan MR, et al. Protection against the Omicron Variant from Previous SARS-CoV-2 Infection. N Engl J Med 2022; 386 (13): 1288–1290.

19. Pulliam JRC, van Schalkwyk C, Govender N, et al. Increased risk of SARS-CoV-2 reinfection associated with emergence of Omicron in South Africa. Science 2022: eabn4947.

20. Ferguson N, Ghani A, Cori A, Hogan A, Hinsley W, Volz E, Imperial College COVID-19 Response Team, “Report 49: Growth, population distribution and immune escape of Omicron in England” (WHO Collaborating Centre for Infectious Disease Modelling, MRC Centre for Global Infectious Disease Analysis, Jameel Institute, Imperial College London, 2021); https://www.imperial.ac.uk/media/imperial-college/medicine/mrc-gida/2021-12-16-COVID19-Report-49.pdf (accessed April 1, 2022).

21. Cohen JI, Burbelo PD. Reinfection With SARS-CoV-2: Implications for Vaccines. Clin Infect Dis 2021; 73 (11): e4223–e4228.

22. World Health Organisation: COVID-19 Weekly Epidemiological Update, Edition 74, published 11 January 2022. https://apps.who.int/iris/bitstream/handle/10665/351044/CoV-weekly-sitrep11Jan22-eng.pdf?sequence=1&isAllowed=y (accessed February 22, 2022).

23. Miljanovic D, Milicevic O, Loncar A, Abazovic D, Despot D, Banko A. The First Molecular Characterization of Serbian SARS-CoV-2 Isolates from a Unique Early Second Wave in Europe. Front Microbiol 2021; 12: 691154.

24. Vidanović D, Tešović B, Volkening J, et al. First whole-genome analysis of the novel coronavirus (SARS-CoV-2) obtained from COVID-19 patients from five districts in Western Serbia. Epidemiol Infect 2021; 149: E246.

25. Slezak J, Bruxvoort K, Fischer H, Broder B, Ackerson B, Tartof S. Rate and severity of suspected SARS-Cov-2 reinfection in a cohort of PCR-positive COVID-19 patients. Clin Microbiol Infect 2021; 27 (12): 1860.e7-1860.e10.

26. Abu-Raddad LJ, Chemaitelly H, Bertollini R. National Study Group for COVID-19 Epidemiology. Severity of SARS-CoV-2 Reinfections as Compared with Primary Infections. N Engl J Med 2021; 385 (26): 2487–2489.

27. Taylor CA, Whitaker M, Anglin O, et al. COVID-19–Associated Hospitalizations Among Adults During SARS-CoV-2 Delta and Omicron Variant Predominance, by Race/Ethnicity and Vaccination Status — COVID-NET, 14 States, July 2021–January 2022. MMWR Morb Mortal Wkly Rep 2022; 71: 466–473.

28. Tso WWY, Kwan M, Wang YL, et al. Intrinsic Severity of SARS-CoV-2 Omicron BA.2 in Uninfected, Unvaccinated Children: A Population-Based, Case-Control Study on Hospital Complications. Lancet 2022; published online March 21. http://dx.doi.org/10.2139/ssrn.4063036 (preprint).

29. Dhillon RA, Qamar MA, Gilani JA, et al. The mystery of COVID-19 reinfections: A global systematic review and meta-analysis. Ann Med Surg (Lond) 2021; 72: 103130.

30. Wang J, Kaperak C, Sato T, Sakuraba A. COVID-19 reinfection: a rapid systematic review of case reports and case series. J Investig Med 2021; 69 (6): 1253–1255.

31. Maier H.E, Balmaseda A, Ojeda S, et al. An immune correlate of SARS-CoV-2 infection and severity of reinfections. medRxiv 2021; published online November 24. https://doi.org/10.1101/2021.11.23.21266767 (preprint).

32. Ioannidis JPA. Factors influencing estimated effectiveness of COVID-19 vaccines in non-randomised studies. BMJ Evid Based Med 2022: bmjebm-2021-111901. https://doi.org/10.1136/bmjebm-2021-111901.

33. Xu S, Huang R, Sy LS, et al. COVID-19 Vaccination and Non-COVID-19 Mortality Risk - Seven Integrated Health Care Organizations, United States, December 14, 2020-July 31, 2021. MMWR Morb Mortal Wkly Rep 2021; 70 (43): 1520–1524.

34. Sciscent BY, Eisele CD, Ho L, King SD, Jain R, Golamari RR. COVID-19 reinfection: the role of natural immunity, vaccines, and variants. J Community Hosp Intern Med Perspect 2021; 11 (6): 733–739.

